# Economic value of resistance-guided gonorrhea treatment: cost-neutrality thresholds for resistance test pricing in the United States

**DOI:** 10.64898/2026.04.07.26350302

**Authors:** Brooke E Nichols, Betsy Wonderly Trainor, Grace Hampson, Yonatan H. Grad, Jeffrey D Klausner

## Abstract

**Background:** Rising antimicrobial resistance in *Neisseria gonorrhoeae* threatens the effectiveness of existing therapies. Resistance-guided treatment (RGT) may reduce treatment failures, complications, and inappropriate use of last-line agents while slowing resistance emergence.

**Methods and Findings:** We developed an individual-level stochastic simulation model of gonorrhea diagnosis and treatment in the United States, incorporating infection prevalence, symptom status, diagnostic accuracy, resistance profiles, treatment pathways, and partner management (costs in 2025 USD). We evaluated three resistance testing strategies, ciprofloxacin-only, ciprofloxacin+ceftriaxone, and triple-target (including a novel drug A), across a wide range of resistance scenarios. We quantified economic value across three dimensions: (1) per-episode direct medical cost savings, (2) system-level costs attributable to ceftriaxone resistance emergence among MSM, and (3) avoided costs of new antibiotic development, estimating the maximum per-test price at which RGT remains cost-neutral. Per-episode cost-neutrality thresholds ranged from near $0 when ceftriaxone resistance was absent to up to $45/test at 15% ceftriaxone resistance. At 50% ciprofloxacin and 5% ceftriaxone resistance, the population-weighted threshold was $4 (95% UI:$3-$8) for a CIP-only test and $11 (95% UI:$5-$14) for a triple-target test. Among MSM, incorporating system-level resistance emergence costs and avoided antibiotic development costs increased the total per-test value to $35–$145 for a single-target test and $84–$128 for a triple-target test, depending on whether prescribing practices shift when ceftriaxone resistance reaches 5%.

**Conclusions:** Resistance-guided therapy offers economic benefits across multiple dimensions even at relatively high diagnostic prices, supporting investment in gonorrhea resistance testing to improve partner outcomes, delay resistance emergence, and enhance the long-term cost-efficiency of gonorrhea management.

## INTRODUCTION

Gonorrhea remains a pressing global public health concern, with an estimated 82 million incident cases annually.^1^ Rising antimicrobial resistance in *Neisseria gonorrhoeae* threatens the effectiveness of ceftriaxone, which is the only remaining recommended empiric therapy.^2,3^ The spread of resistant strains has prompted urgent calls for both novel therapeutics and improved diagnostic strategies to guide tailored treatment. Two new antimicrobials, zoliflodacin and gepotidacin, are starting to enter the market,^4–6^ but preserving their long-term efficacy will be critical to avoid repeating the cycle of rapid resistance emergence that has characterized much of gonorrhea treatment over the past century.^7^

Gonorrhea imposes a significant burden on health systems due to its complications. Among women, untreated or inadequately treated infections can result in pelvic inflammatory disease, chronic pelvic pain, disseminated gonococcal infection, ectopic pregnancy, and tubal infertility.^8,9^ In men, sequelae such as epididymitis and disseminated gonococcal infection similarly contribute to clinical morbidity and downstream healthcare costs.^1,10^ These outcomes carry not only public health consequences but long-term economic consequences for health systems seeking to control sexually transmitted infections.

Resistance-guided treatment (RGT) has been proposed as one strategy to slow the emergence of resistance, preserve the efficacy of both existing and novel therapies, and reduce expenditures associated with treatment failures and complications.^11,12^ This type of approach can also directly improve outcomes for sexual partners by enabling timely and targeted partner treatment. For example, if the index infection is susceptible to ciprofloxacin, a drug that is inexpensive (and less expensive than current partner treatment), widely available, and easily administered, partner treatment can be delivered simply and effectively, often treating two infections at once and preventing onward transmission.^13^

Modelling studies indicate that the use of diagnostics to direct antimicrobial selection, particularly in high-incidence populations such as men who have sex with men, may substantially delay the spread of resistant strains and extend the useful lifespan of current and pipeline drugs.^11,14^ In parallel, economic evaluations highlight that, while upfront diagnostic costs are nontrivial, preventing both treatment failures and gonorrhea sequelae can yield significant value over medium- and long-term horizons.^15,16^ Moreover, by facilitating appropriate partner treatment and averting persistent infection, resistance testing offers an additional pathway for near-term cost-savings.

Despite this growing body of work, relatively few studies have quantified the economic value of resistance testing across the multiple dimensions relevant to adoption decisions. The value of resistance-guided therapy can be understood at three levels: (1) the near-term budget impact, reflecting direct cost savings from avoided repeat visits, more effective partner management, and fewer complications; (2) the economic value of delaying resistance emergence, which preserves the efficacy of existing low-cost antimicrobials and defers the costs associated with treating resistant infections; and (3) the economic value of avoiding or delaying new antibiotic development, which carries substantial research and development costs ultimately borne across the health system. Importantly, quantifying the value associated with each of these dimensions also enables estimation of the maximum diagnostic price at which resistance testing remains cost-neutral or cost-saving, providing an evidence base for pricing, reimbursement, and adoption decisions.

To address this evidence gap, we developed a simulation model of gonorrhea diagnosis and treatment in the United States that incorporates diagnostic performance, resistance-guided therapy, treatment pathways, partner treatment, and complications. By assigning costs to both immediate clinical management and downstream sequelae, we estimated the per-patient cost of care with and without resistance testing across varying resistance scenarios. Using these estimates, we identified cost-neutral price thresholds for resistance diagnostics, first over short-term clinical decision horizons and then through longer-term impacts of slowing resistance emergence and avoiding new antibiotic development.

## Methods

### Model Overview

We developed an individual-level stochastic simulation model in *RStudio (Version 2025.05.0+496)* to capture the costs and outcomes of gonorrhea treatment under different diagnostic strategies. Each simulated encounter began with a patient presenting to care. The patient was assigned an infection and symptom status, and, if infected, a resistance profile to ciprofloxacin, ceftriaxone, and novel drug A (NDA). Patients then followed one of two diagnostic pathways depending on the scenario: syndromic management, or a same-clinic-visit pathogen identification test followed by reflex resistance testing (with either a single, dual, or triple target test). We assumed pathogen identification must occur during the same encounter, because delayed results would typically lead to empiric treatment for symptomatic patients, limiting the clinical utility of resistance results.

Diagnostic outcomes were generated probabilistically based on test sensitivity and specificity, and treatment algorithms reflected clinical guidelines, modified when test results indicated resistance (Figure 1: model diagram; Table 1: key model assumptions). Parameter uncertainty is represented by probability distributions (Table 1). If treatment matched the resistance profile, infections were cured; if not, patients risked persistent infection. Those with ongoing symptoms could re-present for care and receive further diagnostic workup, while asymptomatic individuals were less likely to return. We captured only the probability of return due to untreated (or incorrectly treated) gonorrhea, not due to other possible infections requiring follow-up.

**Figure 1.**
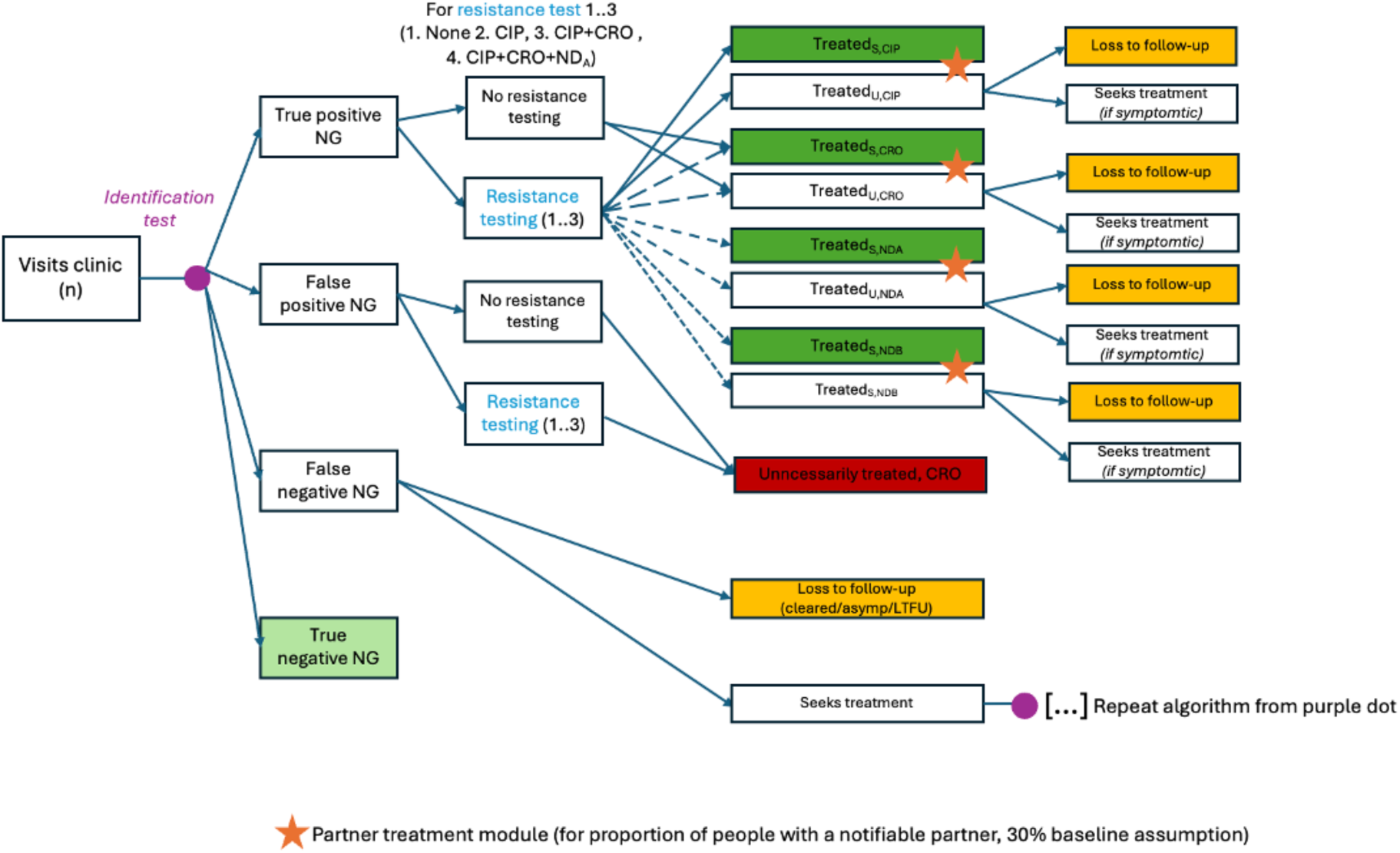
Model diagram: patient pathway

**1.**
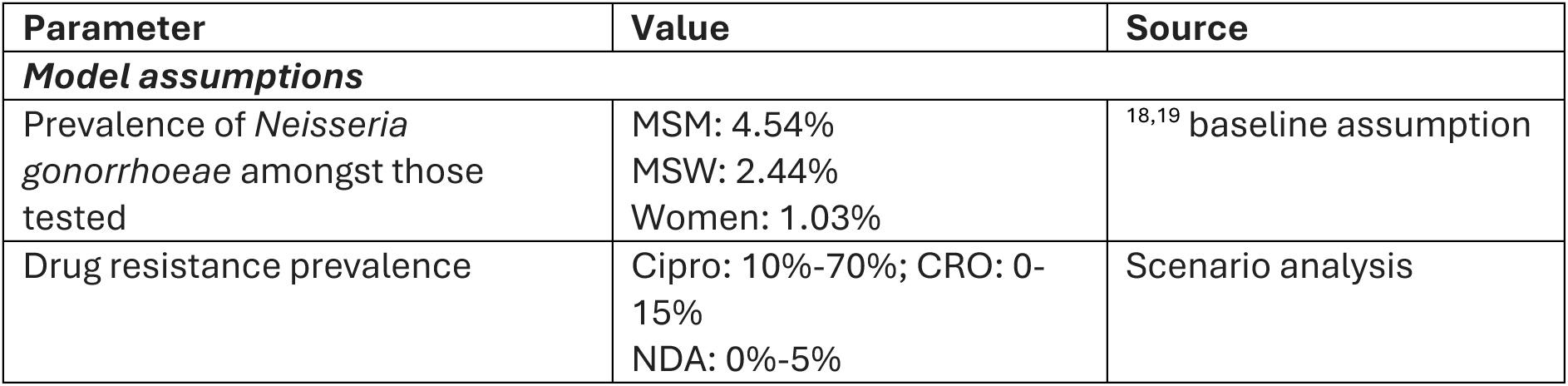
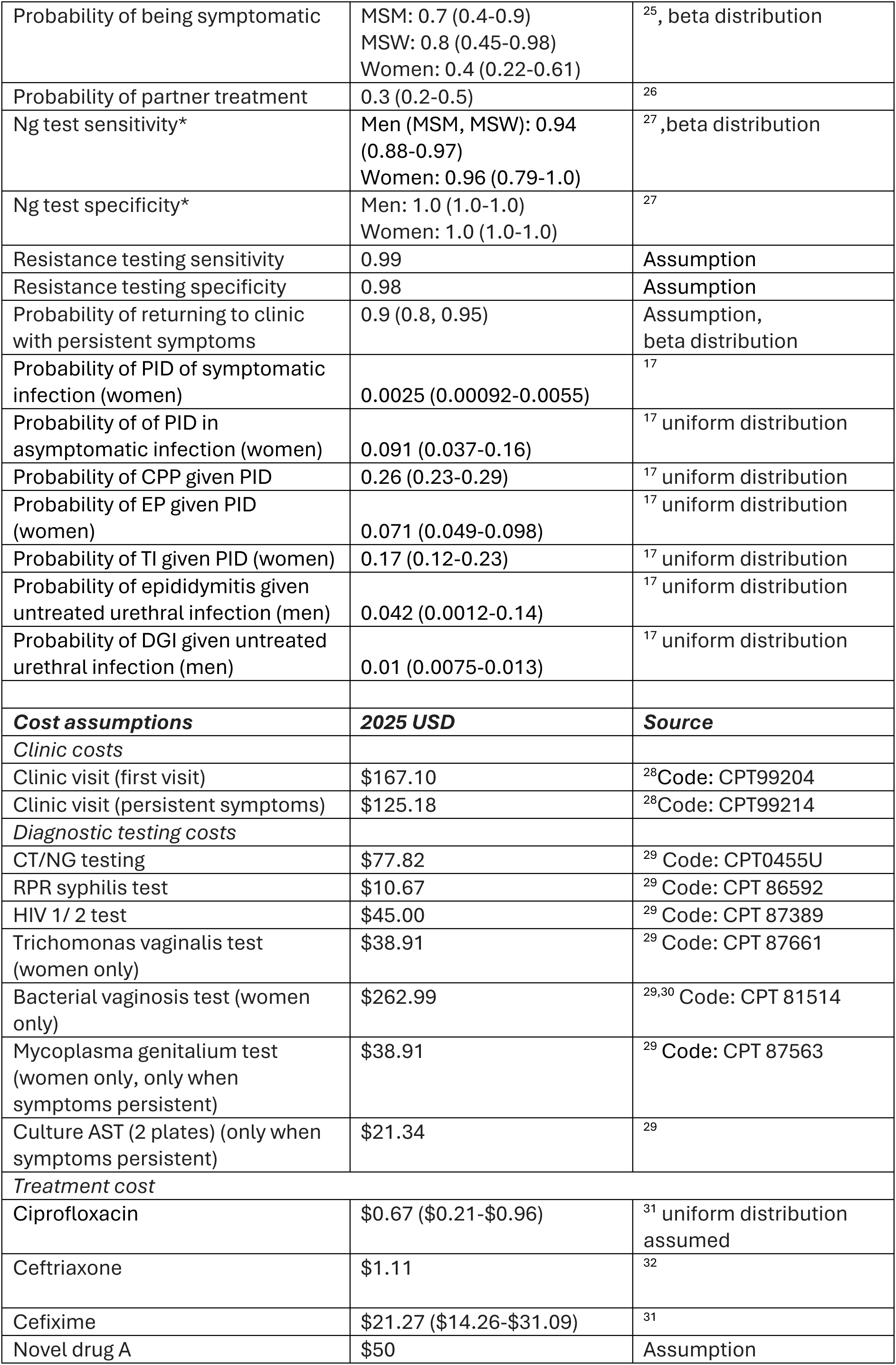
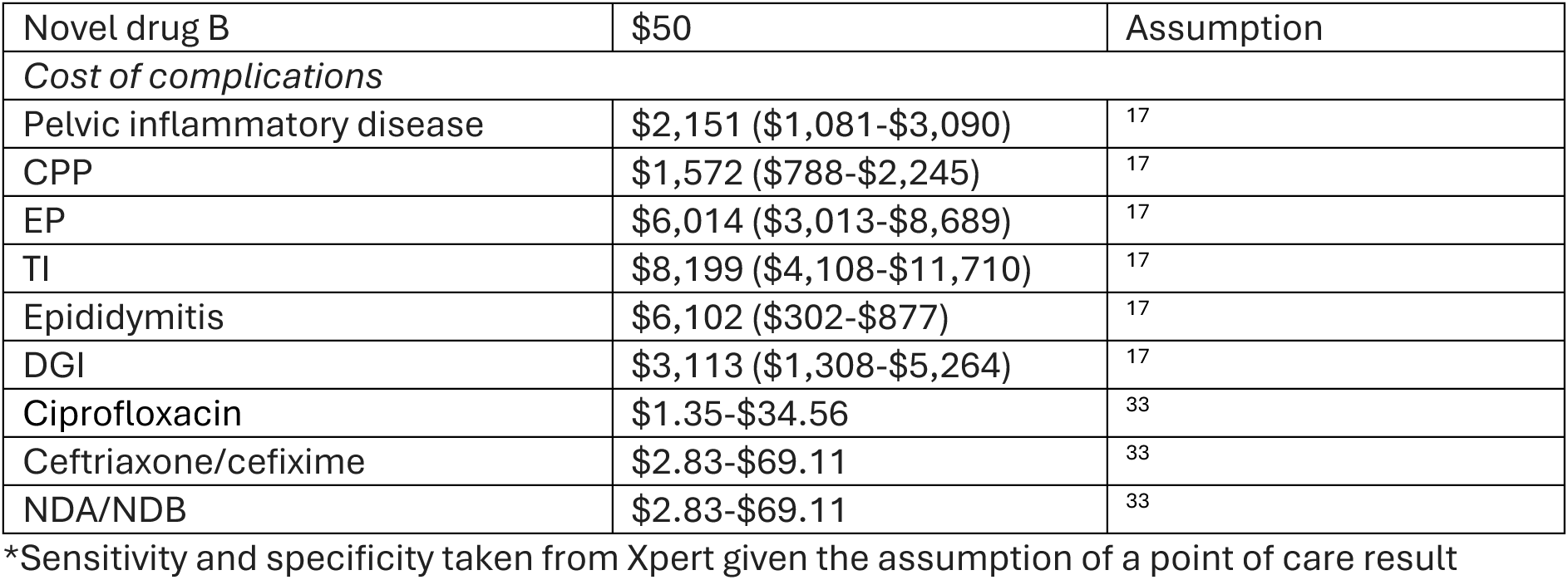
Key model and cost parameters.

Partner treatment was modeled probabilistically, conditional on the index case’s results, symptom status, and treatment pathway. Partners shared the index’s resistance profile and only underwent diagnostic testing if symptomatic after treatment.

The model tracked whether patients achieved cure, remained infected, or developed complications. For women, these included pelvic inflammatory disease (PID) and sequelae such as chronic pelvic pain, ectopic pregnancy, and tubal infertility; for men, complications included epididymitis and disseminated gonococcal infection (DGI), based on U.S. specific estimates.^17^ Costs were assigned at each stage, including clinic visits, diagnostics, drugs, and complication management.^17^ All analyses were conducted using population-specific gonorrhea prevalence among those tested as a baseline (4.5% for men-who-have-sex-with-men [MSM], 2.4% for men-who-have-sex-with-women [MSW], and 1.0% for women).^18,19^ Each scenario (described below) was simulated 10,000 times across 500 probabilistic parameter draws to capture stochastic uncertainty, and results are reported as mean per-patient costs and outcomes with 95% uncertainty intervals, weighted by sex/population group (MSM, MSW, women).

### Resistance Testing Strategies

We compared three hypothetical resistance testing panels against standard diagnostic management without resistance testing. The underlying rationale differs across approaches: ciprofloxacin-only testing is designed to maximize use of ciprofloxacin whenever possible, since it is inexpensive, orally administered, and easily dispensed to partners. Combination panels that include ceftriaxone and novel drug A (NDA) are additionally aimed at ensuring patients receive the most appropriate therapy when multiple resistance strains are present, thereby preserving last-line and emerging agents.

The specific panels evaluated were:

1. CIP-only: testing for ciprofloxacin resistance only
2. CIP+CRO: testing for ciprofloxacin and ceftriaxone resistance
3. CIP+CRO+NDA: testing for ciprofloxacin, ceftriaxone, and a novel agent A (NDA)

Treatment selection followed observed test results, defaulting to guideline-based empiric therapy when no test was available. If no resistance to ciprofloxacin was detected, ciprofloxacin was used first, followed by ceftriaxone, followed by novel drug A and finally novel drug B (if resistance to ciprofloxacin, ceftriaxone, *and* NDA was detected) to which we assume there is no detected resistance across all scenarios.

### Model and cost parameterization

To represent real-world care, we included both syndromic management and diagnostic strategies, with and without resistance testing. Parameters for test sensitivity, specificity, treatment efficacy, partner notification, and complication probabilities were drawn from published literature. Table 1 summarizes all key clinical and cost parameters, along with their sources.

Costs were expressed in 2025 US dollars. We estimated direct medical costs across two components: (1) immediate management costs, including clinic visits, diagnostic testing, antimicrobial treatment, partner treatment, and follow-up care for persistent infection; and (2) the expected costs of complications and sequelae attributable to the index episode, including pelvic inflammatory disease, chronic pelvic pain, ectopic pregnancy, tubal infertility, epididymitis, and disseminated gonococcal infection. Together, these represent the total expected direct medical cost per clinical episode-that is, the full cost consequences of a single gonorrhea testing encounter, including both near-term management and the downstream medical costs of inadequately treated infection.

Drug costs were based on published and publicly available sources; assumptions were made when data were unavailable. Visit and diagnostic costs were derived from US Medicare fee schedules, with appropriate CPT codes identified in consultation with providers and billing specialists.

### Estimation of the value of resistance testing by identifying cost-neutral thresholds

We quantified the economic value of resistance testing across three dimensions, each corresponding to a different scope of cost consequence. For each, we estimated the maximum per-test price at which resistance-guided therapy would remain cost-neutral-that is, the price at which the total costs with resistance testing do not exceed those under standard care.

### 1. Per-episode cost-neutrality

The first dimension captures the value of resistance testing within a single clinical episode. We compared the average per-episode direct medical cost (as defined above) with and without resistance testing across a range of epidemiologic and resistance scenarios. For each population subgroup (MSM, MSW, women), we varied resistance prevalence to ciprofloxacin (10%–70%), ceftriaxone (0%–15%), and novel drug A (0%–5%). For each combination, the model estimated the expected per-patient cost with and without resistance testing, where the cost without testing reflects empiric ceftriaxone-based management.

The cost-neutral threshold price was defined as the maximum diagnostic price at which the average per-patient cost under resistance-guided therapy does not exceed that of standard management. Formally, if *C̄*_0_ is the mean per-patient cost under standard care and *C̄*_*RGT*_is the mean per-patient cost under resistance-guided therapy (with resistance test cost set to zero), then the threshold price per test is:

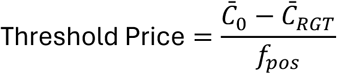

where *f_pos_* is the proportion of individuals testing positive for gonorrhea (and therefore receiving a resistance test). This formulation yields the per-test price at which resistance-guided therapy is cost-neutral when applied to all gonorrhea-positive individuals.

Results were weighted across subgroups by the estimated number of symptomatic and asymptomatic gonorrhea cases among MSM, MSW, and women to derive overall mean threshold prices.^11^

### 2. System-level cost of resistance emergence

The second dimension extends the analysis to incorporate the population-level economic consequences of increasing ceftriaxone resistance over time. Because the per-episode cost of gonorrhea care rises with resistance prevalence (due to higher rates of treatment failure, repeat visits, and complications), delaying resistance emergence has direct economic value.

We estimated this value over a 25-year time horizon. First, we compared the total system cost in a scenario with 0% ceftriaxone resistance to one with 5% ceftriaxone resistance, reflecting the incremental cost attributable to that change (holding ciprofloxacin resistance at 50% and NDA resistance at 0%). We then multiplied the annual number of MSM presenting for gonorrhea testing by the per-episode cost under each resistance scenario and summed over 25 years, discounting future costs at 3% annually. The difference between these two totals represents the aggregate cost attributable to a 5 percentage-point increase in ceftriaxone resistance.

We used previously published transmission modelling of the gonorrhea epidemic among MSM in the United States to estimate the extent to which resistance testing could delay reaching 5% ceftriaxone resistance.^11^ From that work, testing 10% of MSM diagnosed with gonorrhea using a single-target resistance test could delay reaching 5% resistance by 5 years, and testing 50% of MSM diagnosed with gonorrhea using a triple-target resistance test could prevent the emergence of ceftriaxone resistance over the entire modelled period. For the single-target test, we assumed a linear increase from 0% to 5% ceftriaxone resistance over 15 years, followed by continued linear increase thereafter. For the triple-target test, we assumed resistance remains at 0% for the full 25-year horizon. To translate these system-level savings into a per-test value, we divided the total cost attributable to resistance emergence by the total number of resistance tests required over the relevant time horizon.

Finally, we repeated this analysis under an alternative scenario in which, once ceftriaxone resistance reaches 5%, first-line empiric therapy shifts to NDA ($50 per course). This scenario captures a larger cost differential, because the more expensive replacement drug amplifies the per-episode cost consequences of resistance. Given that the underlying transmission modelling is available only for MSM in the United States, these system-level analyses could not be extended to other populations.

### 3. Economic value of avoiding new antibiotic development

The third dimension estimates the economic value of resistance testing in delaying or avoiding the need to develop a new antibiotic for gonorrhea. We assumed a development cost of approximately $1.2 billion per new antibiotic^20^, that the United States represents 44% of the global market for antibiotics^21^, and the proportion of gonorrhea treatment in the United States that occurs among MSM (Supplement S1, 52%). The development cost attributable to gonorrhea treatment among MSM in the United States is therefore $1.2 billion × 44% × 52% = $275 million.

Using the resistance trajectory estimates described above, we calculated the total number of triple-target resistance tests required to prevent the emergence of resistance over a 25-year time horizon. The per-test value of avoiding new antibiotic development was then estimated by dividing the attributable development cost ($227 million) by the total number of tests required.

This analysis applies only to the triple-target test, as only this strategy was projected to fully prevent resistance emergence in the underlying transmission model.^11^

### Uncertainty analysis

Given the wide number of scenarios evaluated and the probabilistic nature of the model, additional uncertainty analysis was limited to deterministic sensitivity analysis by key additional uncertain parameters. This included determining how changing the specific partner-notification rate affected threshold cost (20% to 50%), and the cost of Novel Drug A and B (varying from $25 to $500- as future prices are uncertain), probability of returning to care with symptoms (70%-95%), and resistance test performance (resistance-detection sensitivity 92%-100%; specificity 96%-100%).

## Results

### Baseline costs without resistance testing

We first estimated the average per-person cost of care for individuals diagnosed with gonorrhea under standard practice (without resistance testing). At 50% ciprofloxacin resistance, 0% ceftriaxone resistance, and 0% NDA resistance, the mean per-person cost of care was $361 (95% UI: $321–$412) for MSM, $563 (95% UI: $384–$849) for MSW, and $916 (95% UI: $697–$1,227) for women (WSM), a weighted average of $710 (95% UI: $543–$951) (Figure 2). The higher costs for MSW reflect the greater burden of partner care, while higher costs for women are driven by more extensive diagnostic workup and greater complication costs. These costs reflect the combined costs of initial clinic visits, diagnostics, treatment, partner care, and gonorrhea-related complications. Increasing ceftriaxone resistance increased the average cost of care due to higher rates of incorrect empiric treatment and downstream clinical management; when ceftriaxone resistance increases to 5%, the cost per person with gonorrhea who is tested increases in MSM to $374 (95% UI: $334–$432) and in women to $933 (95% UI: $732–$1,226), and is similar among MSW at $576 (95% UI: $396–$854), with a weighted average of $726 (95% UI: $568–$957) per person diagnosed with gonorrhea. This 5% increase in ceftriaxone resistance alone results in an increase in the total cost of gonorrhea care by 2.2% from $772 million/year to $789 million/year in the United States, when multiplying the annual number of diagnosed and treated cases (Supplement S2.1) by the cost per case. Assuming no shift in first-line antibiotics with an increase in ceftriaxone resistance, the annual cost of 15% resistance to ceftriaxone would cost $824 million annually for gonorrhea-related care in the U.S., a 6.8% increase in total costs.

**Figure 2.**
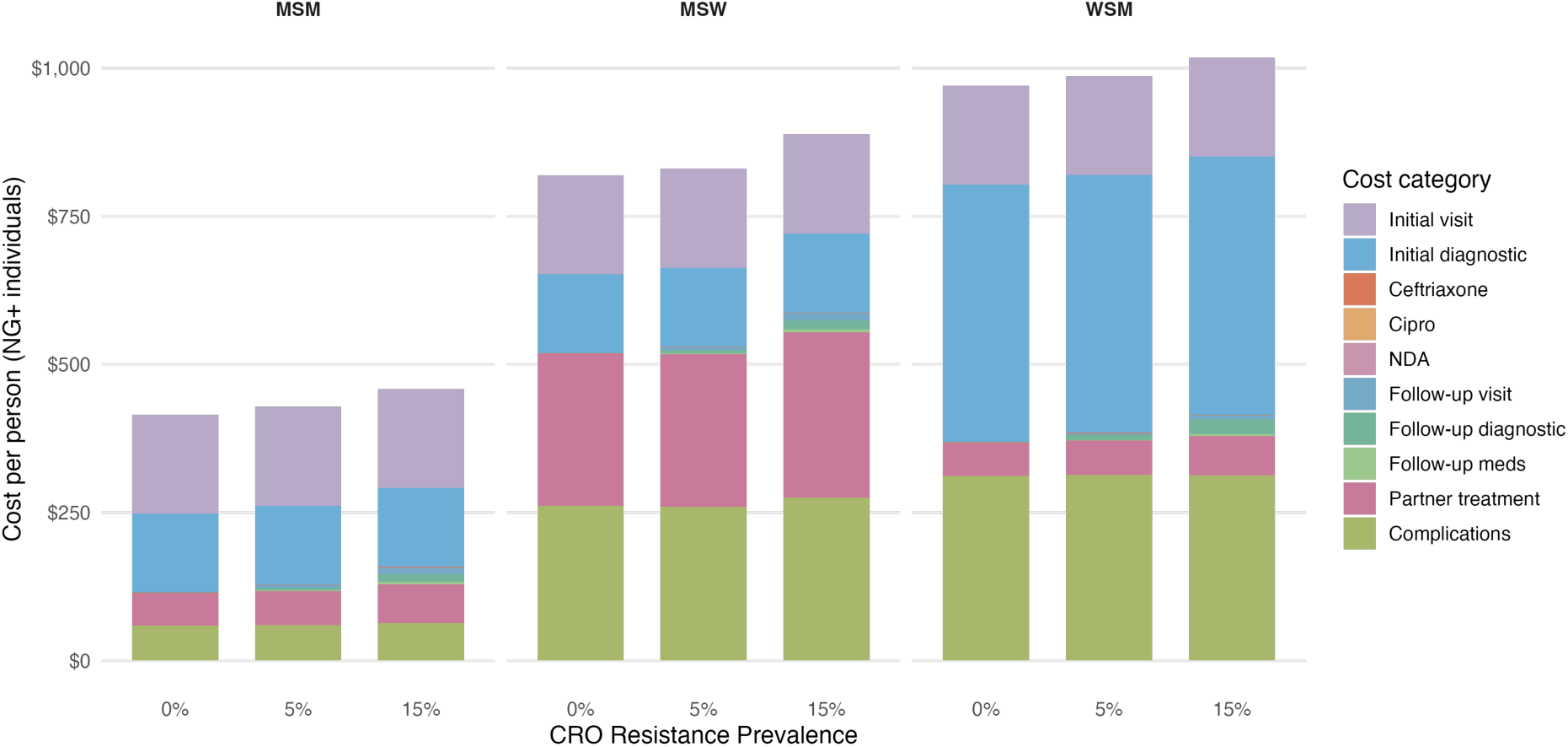
Average per-person cost of gonorrhea care by population group and ceftriaxone resistance, disaggregated by cost category, assuming no resistance testing. The cost of complications includes costs from both own and partner complications; partner complications are weighted by the proportion of individuals who completed partner notification per simulation.

For people diagnosed with gonorrhea, in all scenarios, the cost per person with gonorrhea is largely driven by the initial visit cost (46% of total cost for MSM, 30% for MSW, and 18% for women) and initial diagnostic workup cost (37% of total cost for MSM, 24% for MSW, and 48% for women) (Figure 3). Other key cost drivers included the cost of partner treatment for MSW (46%), and the cost of complications for the index client and their partners (16% of total cost for MSM, 46% for MSW, and 34% for women) (Figure 3). When ceftriaxone resistance is low or absent, resistance testing offers limited savings because empiric ceftriaxone therapy is already effective for most patients. However, once ceftriaxone resistance reaches approximately 5% or higher, the introduction of a CIP-only or CIP+CRO resistance test reduces total per-episode costs relative to standard care. This is because higher resistance prevalence increases the probability of empiric treatment failure, which in turn increases costs from follow-up visits, repeat diagnostics, and complications-costs that resistance-guided therapy can avoid.

**Figure 3.**
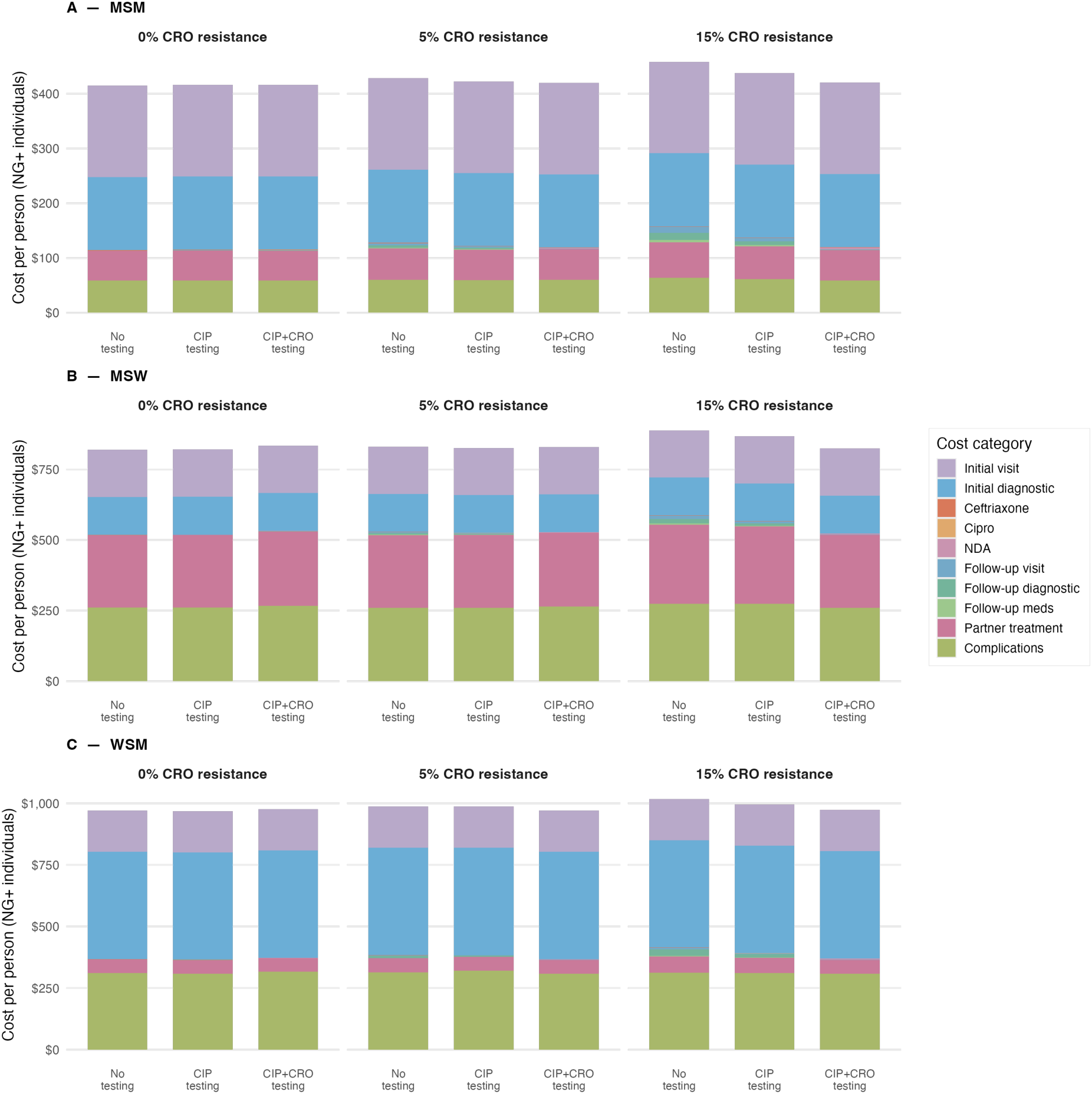
Per-person cost of gonorrhea care across population groups and increasing ceftriaxone resistance, disaggregated by cost category, assuming no resistance testing. The cost of complications includes costs from both own and partner complications; partner complications are weighted by the proportion of individuals who completed partner notification per simulation.

### Estimation of the value of resistance testing by identifying cost-neutral thresholds

#### Short-term cost neutrality estimation

Rising prevalence of ciprofloxacin resistance decreased the cost-neutral threshold for all resistance panels, because fewer patients had ciprofloxacin-susceptible infections, reducing the opportunity to replace ceftriaxone with a lower-cost oral alternative (Figure 4). In other words, the value of a resistance test that identifies ciprofloxacin susceptibility diminishes as the proportion of patients who could benefit from that result shrinks, illustrating that the economic value of a resistance test is inherently linked to the availability and cost of the treatment it enables. Conversely, increasing ceftriaxone resistance raised the per-test cost-neutral price from near $0 (and in some instances <$0, when ceftriaxone resistance was 0%) to as high as $45 per test when ceftriaxone resistance reached 15%. This occurred because higher ceftriaxone resistance increased the probability of empiric treatment failure under standard care, which in turn increased costs from repeat visits, diagnostic workup, partner care, and complications.

**Figure 4.**
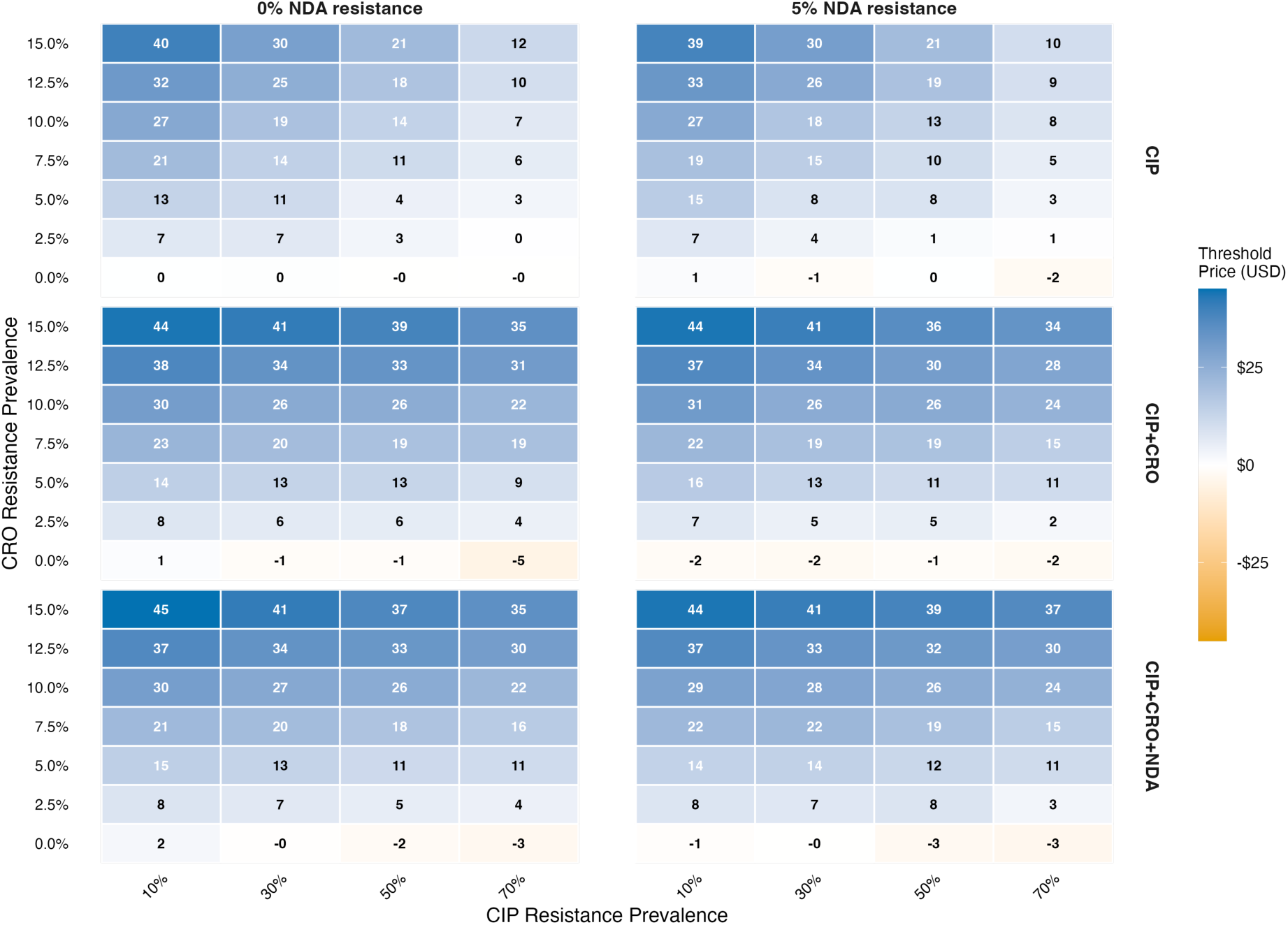
Cost neutral threshold price for a reflex Ng resistance test (CIP-only, CIP+CRO, CIP+CRO+NDA), or the price for a reflex resistance test at which no additional costs to the healthcare system would be incurred

Across scenarios, cost-neutrality thresholds for resistance tests varied widely, and estimates were also subject to stochastic variability. At 50% ciprofloxacin resistance, 5% ceftriaxone resistance, and 0% novel drug A resistance, the population-weighted cost-neutral price per CIP-only resistance test was $4 (95% UI $3-$8), and $11 (95% UI $5-$14) for a triple-target (CIP+CRO+NDA) test. If the resistance rate to novel drug A is also 5%, then the population-weighted cost-neutral price per CIP-only resistance test is $8 (95% UI $7-$13), and a triple-target resistance test is $12 (95% UI $4-$13).

### System-level cost of resistance emergence (MSM only)

Separately from the per-episode analysis above, we estimated the additional per-test value of using resistance testing to delay increases in ceftriaxone resistance among MSM over a 25-year time horizon. This value represents a distinct economic dimension, or the system-level costs that resistance testing could help avoid and is additive to the per-episode cost-neutrality thresholds reported above.

Under a scenario in which prescribing practices do not change when ceftriaxone resistance emerges, the additional per-test value of delaying resistance is $29 (95% UI $27-$35) for a CIP-only test and $22 (95% UI $20-$27) for a triple-target test. If instead first-line therapy transitions to a new antibiotic (assumed cost of $50/course) once ceftriaxone resistance reaches 5%, the additional per-test value increases to $139 (95% UI $49-$206) for a CIP-only test (applied to 10% of diagnosed cases) and $66 (95% UI $32-$93) for a triple-target test (applied to 50% of diagnosed cases).

### Economic value of avoiding new antibiotic development (MSM only)

As a further additional dimension, we estimated the per-test value of avoiding or delaying the need to develop a new antibiotic for gonorrhea. If a triple-target test were applied to 50% of all MSM diagnosed with gonorrhea, approximately 6.2 million tests (range 5.2–7.8 million) would be required to prevent an increase in resistance over the 25-year time horizon. Dividing the attributable development cost ($275 million) by this number of tests yields an additional per-test value of $51 (UR $41–$61).

### Combined per-test value across all three dimensions

Summing across all three dimensions- per-episode savings, resistance emergence costs, and avoided drug development- the total cost-neutral per-test value of a single-target resistance test ranges from $35 ($33-$42) to $145 ($55-$213), depending on whether prescribing practices change when ceftriaxone resistance reaches 5%. For a triple-target resistance test, the total cost-neutral per-test value ranges from $84 ($71-$100) to $128 ($83-$166) (Table 2).

**Table 2.** Per-test value of a single-target resistance test and a triple-target resistance test in MSM when 50% cipro resistance, 5% ceftriaxone resistance, and 0% resistance to novel antibiotic A.

|  | Mean per-test value to reach cost-neutrality (single-target test) | Mean per-test value to reach cost-neutrality (triple-target test)* |
| --- | --- | --- |
| Short term to healthcare system (A) | \$6 (\$6-\$7) | \$11 (\$10-\$12) |
| Long term if the use of testing results in reductions in overall resistance rates (B) | \$29 (\$27-\$35) | \$22 (\$20-\$27) |
| Long term if the use of testing results in reductions in overall resistance rates (if transitioning to a new antibiotic at 5% resistance to ceftriaxone) (C) | \$139 (\$49-\$206) | \$66 (\$32-\$93) |
| Long term avoiding new antibiotic development (D) | N/A* | \$51 (\$41-\$61) |
| TOTAL (A+B+D) | \$35 (\$33-\$42) | \$84 (\$71-\$100) |
| TOTAL (A+C+D) | \$145 (\$55-\$213) | \$128 (\$83-\$166) |

\*per-test value is lower compared to single-target test as more tests are required to fully reduce increases in resistance (50% as compared to 10%)

## Discussion

In this study, we developed a simulation model of gonorrhea diagnosis and treatment in the United States that incorporates diagnostic accuracy, resistance-guided therapy, partner treatment, and the probability of complications. We quantified the value of resistance testing by estimating the diagnostic price at which resistance-guided therapy remains cost-neutral compared with standard care. Across resistance scenarios and testing strategies, resistance testing generated value through several mechanisms: by substituting lower-cost antimicrobials when efficacious, by reducing repeat visits and follow-up diagnostics associated with persistent infection, and by preventing costly complications among index cases and partners. Importantly, the value of resistance testing extended beyond short-term care episodes. When accounting for the downstream costs of increasing ceftriaxone resistance, and the potential to delay the need for new antibiotic development, cost-neutral thresholds increased substantially, particularly among MSM.

The dimension of economic value considered had a major impact on the estimated per-test value. When considering only the near-term budget impact—direct cost savings from avoided repeat visits, more effective treatment, and fewer complications—the cost-neutral threshold was modest. Incorporating the economic value of delaying resistance emergence and the costs of new antibiotic development substantially increased the estimated per-test value. Notably, the per-test value of a multi-target test was lower than that of a single-target test in some scenarios because achieving the larger long-term benefit required using the test more broadly. This illustrates that test value depends not only on performance characteristics, but also on strategic deployment within a population and the anticipated evolution of resistance.

These findings have implications for diagnostic developers, payers, and guideline-setting bodies. First, the cost-neutral thresholds identified in this study indicate that gonorrhea resistance assays could be priced substantially higher than current molecular diagnostics and still be cost-neutral or cost-saving, even when only per-episode direct medical costs are considered. When the system-level value of delaying resistance and avoiding new antibiotic development is included, the economic case strengthens further. This said, this additional headroom is created because of the test and treatment in combination and further thinking and analysis is required to establish the appropriate attribution of value across these two technologies. Second, resistance-guided therapy generates economic benefits for partners as well as index patients, reflecting a broader programmatic value than is typically captured in diagnostic evaluations. Finally, by quantifying each dimension of value separately—per-episode costs, resistance emergence, and drug development—this analysis allows decision-makers to weigh the components most relevant to their context, whether that is near-term episode-level costs or longer-term system-level consequences.

Several studies have evaluated the clinical, epidemiological, and programmatic value of resistance-guided gonorrhea therapy, but relatively few have focused explicitly on quantifying the economic value of resistance testing across multiple dimensions, from per-episode direct medical costs through to the system-level value of preserving antimicrobial efficacy and avoiding new drug development costs. Prior modelling and clinical work indicate that resistance-guided and susceptibility-guided strategies can preserve the effectiveness of ceftriaxone and other current or pipeline agents by distributing selective pressures across multiple antibiotics, particularly in populations such as MSM and in settings with heterogeneous resistance prevalence. The present results are consistent with this evidence base and further explicitly incorporate partner treatment and complications into the short-term analyses.^16,22,23^ Clinical and economic studies of *gyrA*-based testing further suggest that using ciprofloxacin when susceptibility is known is highly efficacious and can be cost-saving or cost-neutral relative to guideline-recommended empiric therapy, especially in higher-incidence populations. The current analysis extends these findings by showing that cost-neutral or cost-saving thresholds for resistance-guided strategies remain favorable across a wide range of resistance prevalence scenarios and when multi-target assays are considered.^22,23^ Economic evaluations of STI diagnostics have highlighted the long-term value of more accurate or rapid tests in reducing treatment failures, sequelae, and onward transmission, although few have explicitly quantified diagnostic price thresholds for gonorrhea resistance testing. By estimating per-test values under both short- and long-term perspectives, this work provides a complementary framework to existing diagnostic and resistance-guided treatment studies and contributes evidence that is directly relevant for payers and health systems considering adoption of such assays.

This analysis has several limitations. First, we assumed that clinical care follows recommended guidelines. In practice, treatment may diverge from guidelines due to patient preference, clinical judgment, or barriers to treatment access. If real-world management incurs lower costs due to less intensive care pathways, the per-test value of resistance testing may be reduced.

Second, our model assumes that resistance results, however they are ultimately generated (same-visit or centralized), are used to guide treatment selection. In practice, most pathogen identification testing is currently laboratory-based, and empiric treatment may still be initiated for symptomatic patients before results are available. This could reduce the immediate treatment benefits for symptomatic patients in some settings. However, resistance information returned later would still influence management for asymptomatic individuals and partners, and therefore many of the downstream cost and complication savings we estimate would remain. Importantly, the cost-neutral thresholds reported here reflect the maximum price of the resistance test itself, and do not include associated operational costs such as specimen handling, laboratory processing, or additional clinician time. The total cost of implementing resistance testing in each workflow may therefore be higher than the test price alone, and the thresholds we report should be interpreted as an upper bound on the test price component within that total cost. Given that the prevalence of gonorrhea among all tested is relatively low at the national level,^19^ we expect our cost-neutral thresholds to remain stable across different diagnostic workflows.

Third, the long-term economic analyses assume constant annual testing volume and case counts over time. These analyses were designed to illustrate the potential cost attributable to increasing resistance and the value of delaying or preventing that increase, rather than to forecast the trajectory of the gonorrhea epidemic or population growth.

Fourth, outcomes were based on direct medical costs and did not include indirect costs, productivity losses, or broader social impacts. Incorporation of these effects would likely increase the value of resistance-guided therapy. Indeed, steps are being taken to explore how some of these additional elements can be captured in economic analyses, for example via the STRIDES conceptual framework.^24^ The framework introduces seven AMR-specific value elements that are tailored to the unique role diagnostics play in preserving antibiotic effectiveness and projecting public health. Work is ongoing to explore how this conceptual framework can be operationalised within economic analysis, allowing the full value of these diagnostics to be captured.

Fifth, long-term economic estimates were restricted to MSM because the underlying modelling work quantifying resistance trajectories is available only for MSM in the United States.^11^ Given the epidemiology of gonorrhea in MSM, these estimates likely represent an upper bound on per-test value, and similar analyses in other populations may yield somewhat lower values. Finally, transmission dynamics were not explicitly modeled, which means that the population-level benefits of reducing resistance prevalence are likely underestimated. For simplicity, we evaluated scenarios in which resistance prevalence and associated costs were set at fixed values rather than evolving over time. This allowed us to compare clinical and economic outcomes across different “worlds” of resistance but does not capture how resistance might spread or decline in response to treatment strategies.

Despite these limitations, this study demonstrates the economic value of resistance testing for gonorrhea across multiple dimensions. Even when considering only per-episode direct medical costs, resistance-guided therapy can reduce treatment failures, prevent complications, and improve partner management. When the system-level consequences of resistance emergence and the costs of new antibiotic development are also considered, the economic case for resistance testing strengthens substantially. Together, these findings suggest that resistance testing offers a pathway to more efficient use of existing antimicrobials and more sustainable management of antimicrobial resistance. As novel diagnostics and resistance assays become increasingly available, the framework presented here, which separately quantifies per-episode, resistance-emergence, and drug-development value, can support pricing, reimbursement, and implementation decisions in both clinical and public health settings.

## Data Availability

All data produced in the present work are contained in the manuscript

## SUPPLEMENT

S1. Model description and equations

S2. Calculations for total diagnosed cases and number of tests

S3. Deterministic sensitivity analysis

## Acknowledgements

This work was supported by CARB-X, which is funded in part with federal funds from the US Department of Health and Human Services (HHS), Administration for Strategic Preparedness and Response, Biomedical Advanced Research and Development Authority (BARDA) under agreement number 75A50122C00028, and by awards from Wellcome (WT224842), the UK Department of Health and Social Care’s Global Antimicrobial Resistance Innovation Fund (GAMRIF), the Gates Foundation, Germany’s Federal Ministry of Research, Technology and Space (BMFTR), the Novo Nordisk Foundation, Italy’s Ministry of Economy and Finance (MEF), Japan’s Ministry of Health, the European Commission’s DG Health Emergency Preparedness and Response Authority (DG HERA), and KfW Development Bank. NIAID, part of the NIH, provides support in the form of in-kind services through access to a suite of preclinical services for product development. The content of this manuscript is solely the responsibility of the authors and does not necessarily represent the official views of any CARB-X funders.

## Notes

### Competing Interest Statement

The authors have declared no competing interest.

