## Supplemental information for "Economic value of resistance-guided gonorrhea treatment: cost-neutrality thresholds for resistance test pricing in the United States"

### S1. Model Description and Equations

This appendix provides the mathematical formulation underlying the individual-level stochastic simulation model described in the main text. The model simulates clinical encounters for individuals presenting for gonorrhea testing and tracks costs across diagnostic, treatment, partner management, and complication pathways.

#### S1.1 Population and Infection Assignment

Each simulated individual i in population group $g \in$ {MSM, MSW, WSM} is assigned:

> *Infection status:* $NG_i \sim \text{Bernoulli}(\pi^g)$, where $\pi^g$ is the prevalence of *N. gonorrhoeae* among those tested in group g ($\pi_{MSM} = 0.0454$; $\pi_{MSW} = 0.0244$; $\pi_{WSM} = 0.0103$).[1]
>
> *Symptom status:* $S_i \sim \text{Bernoulli}(p_{sympt,g})$, where $p_{sympt,g} \sim \text{Beta}(\alpha^g, \beta^g)$ as specified in Table 1.
>
> *Resistance profile* (conditional on infection): For each antibiotic $a \in$ {CIP, CRO, NDA}, $R_{i,a} \sim \text{Bernoulli}(r_a)$, where $r_a$ is the prevalence of resistance to antibiotic a, varied across scenario analyses.

#### S1.2 Diagnostic Testing

All individuals undergo gonorrhea diagnostic testing. The test result $T_i$ is generated as:

$$T_i = 1 \text{ with probability: } Se^g \text{ if } NG_i = 1, \text{ or } (1 - Sp^g) \text{ if } NG_i = 0$$

where $Se^g$ and $Sp^g$ are the group-specific sensitivity and specificity of the gonorrhea NAAT. For men (MSM, MSW): Se ~ Beta(99.64, 6.36), Sp = 1.0. For women (WSM): Se ~ Beta(11.89, 0.50), Sp = 1.0.[2]

For individuals testing positive ($T_i = 1$), resistance test results are generated for each antibiotic a included in the resistance panel:

$$RT_{i,a} = 1 \text{ with probability: } Se_{ra} \text{ if } R_{i,a} = 1, \text{ or } (1 - Sp_{ra}) \text{ if } R_{i,a} = 0$$

where $Se_{ra} = 0.99$ and $Sp_{ra} = 0.98$ for all antibiotics tested.

#### S1.3 Treatment Selection

Treatment is assigned hierarchically based on resistance test results. The preferred drug order is: ciprofloxacin (CIP) > ceftriaxone (CRO) > novel drug A (NDA) > novel drug B (NDB).

> **Without resistance testing (standard care):** All test-positive individuals receive ceftriaxone as empiric therapy.
>
> **With resistance testing:** The first-line drug is the highest-preference antibiotic for which the resistance test result indicates susceptibility (i.e., test result negative for resistance). If all tested drugs show resistance, a fallback drug is used: CRO for CIP-only panels, NDA for CIP+CRO panels, and NDB for triple panels.

Treatment success is defined as the administered drug matching the true susceptibility profile (i.e., the individual is not truly resistant to the administered drug). NDB is assumed universally effective across all scenarios.

#### S1.4 Follow-up and Re-treatment

If treatment fails (drug administered to a truly resistant infection) AND the individual is symptomatic AND returns to care:

P(return | symptomatic, treatment failure) ~ Beta(54.45, 6.05), mean ≈ 0.90 (assumption)

Returning individuals incur a follow-up visit cost, follow-up diagnostic workup, and receive a second-line drug selected using the same hierarchical algorithm, advancing to the next available drug.

Asymptomatic individuals with failed treatment do not return for further care in the model and remain at risk for complications.

#### S1.5 Partner Treatment

Partner engagement occurs with probability $p_{partner}$ ~ Beta(10.47, 24.43), mean ≈ 0.30.[3] When a partner is engaged:

(1) The partner receives the same first-line drug as the index case.

(2) If the index case is truly infected, the partner is assumed to share the same resistance profile.

(3) Partner treatment failure follows the same logic as index re-treatment (conditional on partner symptoms).

(4) Partner complications are modeled identically to index complications, with sex-specific probabilities based on the partner's sex.

(5) Partners of false-positive index cases receive treatment but incur no complication costs, as no true infection is present.

#### S1.6 Complications

**Women (index or partner):** PID probability depends on symptom status: p(PID | symptomatic) ~ U(0.00092, 0.0055); p(PID | asymptomatic) ~ U(0.037, 0.16). Importantly, PID risk accrues regardless of treatment success, reflecting the biological lag between infection and sequelae. Conditional on PID: p(CPP|PID) ~ U(0.23, 0.29); p(EP|PID) ~ U(0.049, 0.098); p(TI|PID) ~ U(0.12, 0.23). Total complication cost = $C_{PID}$ + p(CPP|PID)×$C_{CPP}$ + p(EP|PID)×$C_{EP}$ + p(TI|PID)×$C_{TI}$.[4]

**Men (index or partner, if untreated):** Complication cost = p(epididymitis)×$C_{epi}$ + p(DGI)×$C_{DGI}$, where p(epididymitis) ~ U(0.0012, 0.14) and p(DGI) ~ U(0.0075, 0.013).[4] Male complications accrue only if the individual was not successfully treated.

#### S1.7 Cost Accounting

The total cost for individual i is:

$$C_i = C_{init\_visit} + C_{init\_diag} + C_{drug\_1} + C_{fu\_visit} + C_{fu\_diag} + C_{drug\_2} + C_{res\_test} + C_{partner} + C_{complications}$$

where each component is conditional on the pathway taken (see Table 1 in main text for costs).

#### S1.8 Cost-Neutrality Threshold Calculation

The resistance test threshold price is calculated as:

$$\text{Threshold Price} = (\overline{C_0} - \overline{C_{RGT}}) / f_{pos}$$

where $\overline{C_0}$ is the mean per-patient cost under standard care (no resistance testing), $\overline{C_{RGT}}$ is the mean per-patient cost under resistance-guided therapy (with test cost set to \$0), and $f_{pos}$ is the fraction of individuals testing positive for gonorrhea (who would receive a resistance test). This yields the maximum per-test price at which RGT is cost-neutral relative to standard care.

#### S1.9 Probabilistic Sensitivity Analysis

For each scenario (group × CIP resistance × CRO resistance × panel), 1,000 probabilistic parameter draws were performed. Each draw sampled all uncertain parameters from their respective distributions (Table 1). Within each draw, 20,000 individuals were simulated. Results are reported as means with 95% uncertainty intervals (2.5th and 97.5th percentiles across the 1,000 PSA draws).

### S2. Calculations for Total Diagnosed Cases and Number of Tests

We estimated the annual number of gonorrhea NAAT tests, test positivity, and diagnosed cases by population group using *Tao & Gift*., [1] which reported test volumes and positivity rates from a nationally representative sample of clinical laboratories.

*Tao & Gift* reported data from laboratories representing an estimated 20–30% of national gonorrhea NAAT testing volume.[5] We scaled reported test counts to national estimates by dividing by the midpoint assumption of 25% (range: 20–30%), yielding a plausible range for total national annual testing volume.

#### *S2.1 Estimated Annual Gonorrhea Tests and Diagnoses*

Table S1 presents estimated annual gonorrhea NAAT test volumes, positivity rates, and diagnosed case counts by population group.[1,5]

**Table S1. Estimated annual gonorrhea NAAT tests, positivity, and diagnosed cases by population group.**

| Group | Annual NAAT tests (national estimate)[1,5] | Gonorrhea positivity (weighted average across sites)[1,6] | Annual diagnosed & treated cases (calculated) |
|---|---|---|---|
| MSM | 11,002,688 (9,168,907–13,753,360) | 4.5% | 499,522 (416,268-624,403) |
| MSW | 8,372,504 (6,977,087–10,465,630) | 2.4% | 204,289 (170,241-255,361) |
| Women | 24,373,581 (20,311,317–30,466,976) | 1.0% | 251,383 (209,486-314,228) |
| **Total** | **43,748,773** | — | **955,194 (795,995-1,193,992)** |

#### *S2.2 Number of Resistance Tests by Strategy*

We estimated the number of gonorrhea resistance tests required annually and over a 25-year horizon under two testing strategies applied to MSM diagnosed with gonorrhea. As explained in the main text, the single-target test, if targeted to 10% of MSM diagnosed with gonorrhea, could delay reaching 5% resistance by 5 years, and the triple-target test, if targetd to 50% of MSM diagnosed with gonorrhea, could prevent the emergence of ceftriaxone resistance across the entire modelled period.[7]

For the system-level analysis, the relevant denominator is the number of NAAT-positive MSM among all those tested nationally- the population who would receive a resistance test at their gonorrhea testing visit. Using an estimated annual MSM NAAT testing volume of 11,002,688 (range 9,168,907–13,753,360, from above) and a gonorrhea positivity rate of 4.5%:

> Single-target (CIP-only), 10% of NAAT-positive MSM: 0.10 × (0.045 × 11,002,688) = 49,952 tests/year → 1,248,805 tests over 25 years (range 1,040,671-1,561,006)
>
> Triple-target (CIP+CRO+NDA), 50% of NAAT-positive MSM: 0.50 × (0.045 × 11,002,688) = 249,761 tests/year → 6,244,025 tests over 25 years (range 5,203,355–7,805,032) based on the testing volume range of 9,168,907–13,753,360)

For context, 49,952 annual resistance tests for the single-target per-episode strategy represent approximately 0.5% of the estimated 11.0 million annual gonorrhea NAAT tests performed among MSM nationally. The 216,203 annual tests for the triple-target per-episode strategy represent approximately 2.0% of annual MSM NAAT volume.

#### S3. Deterministic Sensitivity Analysis

In addition to the probabilistic sensitivity analysis conducted across all scenarios, we performed one-way deterministic sensitivity analyses on the following parameters at a reference scenario of 50% CIP resistance, 5% CRO resistance, and 0% NDA resistance among MSM:

| Parameter | Base case | Range tested | Rationale |
|---|---|---|---|
| Partner notification rate | 30% | 20%–50% | Varies widely by setting |
| Cost of NDA and NDB | $50 | $25–$500 | Future pricing uncertain |
| Probability of return with symptoms | 90% | 70%–95% | Limited data on return rates |
| Resistance test sensitivity | 99% | 92%–100% | Assay-dependent |
| Resistance test specificity | 98% | 96%–100% | Assay-dependent |

**Figure S3. Deterministic Sensitivity Analysis — Tornado Diagrams**

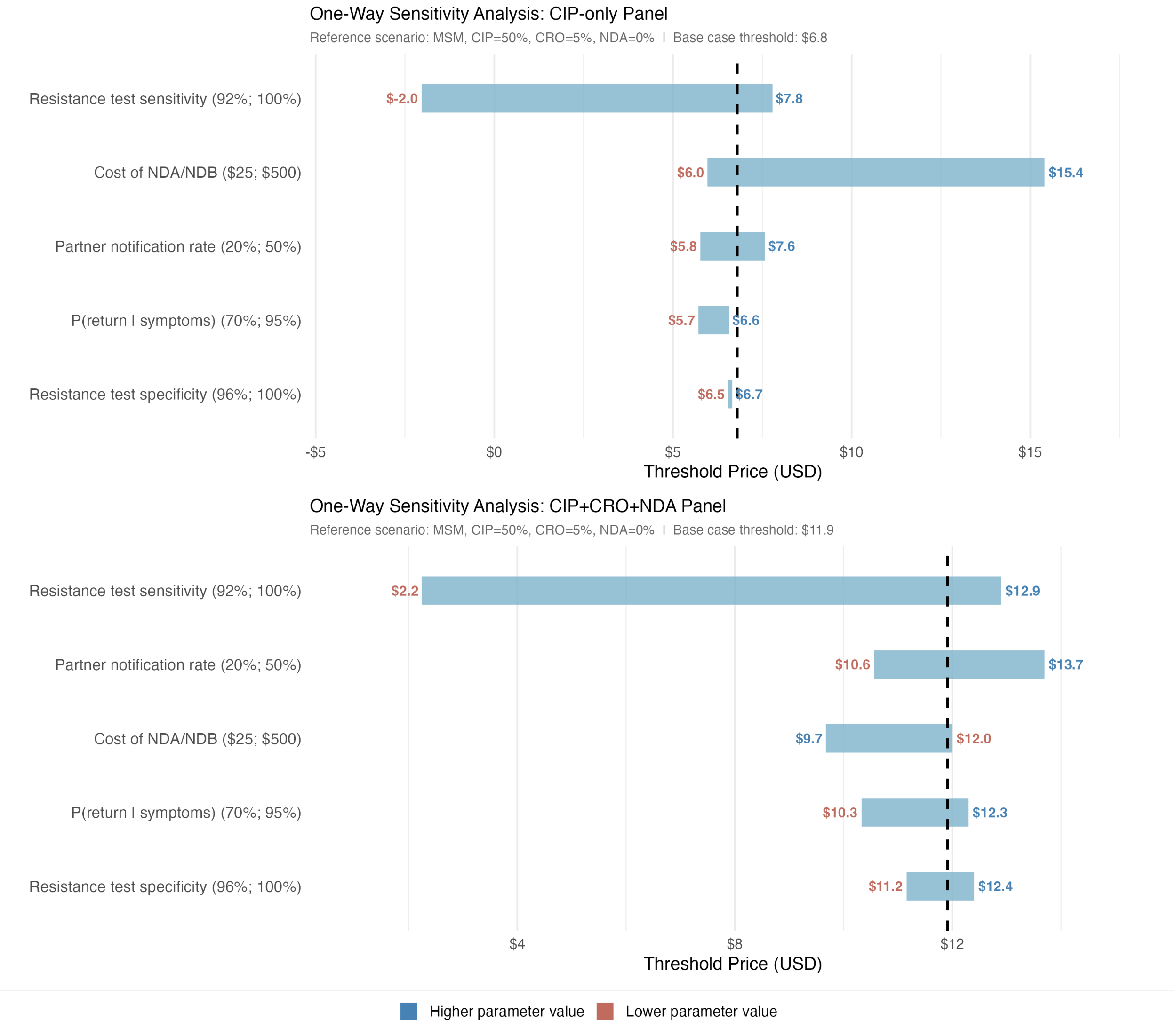


For the CIP-only panel, *resistance test sensitivity* was the most influential parameter, with the threshold ranging from −$2.0 (at 92% sensitivity) to $7.8 (at 100% sensitivity). A low sensitivity means that a substantial proportion of ciprofloxacin-susceptible infections are misclassified as resistant, causing the test to steer patients toward more expensive second-line drugs and eroding the cost savings that would otherwise justify the test. When sensitivity approaches 100%, the test performs near-perfectly and the threshold rises toward its theoretical maximum. *Cost of NDA/NDB* was the second most influential parameter (threshold range $6.0–$15.4): higher drug costs increase the savings generated by correctly avoiding unnecessary escalation to expensive agents, raising the threshold price a CIP-only test can support. *Partner notification rate* (range $5.8–$7.6) and *probability of return given symptoms* (range $5.7–$6.6) had moderate influence, as both parameters affect the volume of follow-up care costs that resistance-guided treatment can avoid. *Resistance test specificity* (range $6.5–$6.8) had the smallest effect, reflecting that at a specificity of 96–100%, the rate of false-positive resistance calls is already low and further improvements yield little additional economic benefit.

For the triple-target (CIP+CRO+NDA) panel, the pattern of influential parameters was broadly similar but with some differences in magnitude and ordering. *Resistance test sensitivity* again produced the widest range ($2.0–$12.9), reflecting its fundamental importance: a triple-target test that frequently misclassifies resistance to any of the three drugs risks steering patients toward inappropriate and potentially expensive treatment cascades. *Partner notification rate* was the second most influential parameter for the triple-target panel (range $10.6–$13.7), a larger relative effect than for the CIP-only panel, likely because the triple-target strategy is deployed more broadly (50% of cases) and therefore partner treatment pathways represent a larger share of total costs. *Cost of NDA/NDB* showed an inverted direction for the triple-target panel (range $9.7–$12.0, higher cost associated with lower threshold), because the triple-target test actively directs CIP- and CRO-resistant patients to NDA as first-line therapy; when NDA is expensive, this increases costs in the with-test arm more than in the no-test arm, reducing the net savings and therefore lowering the threshold. *Probability of return given symptoms* (range $10.3–$12.5) and *resistance test specificity* (range $11.2–$12.4) had the smallest influence on the triple-target threshold.

Across both panels, resistance test sensitivity was consistently the dominant driver of threshold variability, underscoring the importance of assay performance characteristics in determining the economic viability of resistance-guided gonorrhea treatment. All other parameters produced threshold ranges that remained above zero, indicating that the cost-neutrality finding is robust to plausible variation in partner notification rates, return-to-care probabilities, drug costs, and test specificity. The one exception is the CIP-only threshold at the lowest tested sensitivity (92%), which became negative, meaning that a test with this level of sensitivity would not be cost-neutral at any price under the reference scenario.
